# Effect of Pre-Pubertal Body Mass Index Status on Longitudinal Height Trajectory in Vietnamese Children: 2018-2025 School Health Analysis

**DOI:** 10.64898/2026.07.26.26358003

**Authors:** Nhan T. Ho, Michelle Hermiston, Quynh T. Nguyen, Minh A. Nguyen, Quyet V. Nguyen, Anh Q. Dao, Chi T. L. Tran, Quang N. Nguyen, Lam N. Phung, Cuong T. Do, An N. Pham

## Abstract

**Background:** Vietnam and similar low-middle-income countries face a double/triple burden of overweight-obesity, thinness, and stunting. We examined how pre-pubertal BMI affects longitudinal height trajectories in a large Vietnamese cohort.

**Methods:** We analyzed annual school health data from Hanoi, Ho Chi Minh City, and Haiphong (2018–2025). Children with ≥3 visits (n = 40,887) were classified by WHO BMI category (thinness, normal, overweight, obesity) at sex-specific pre-pubertal index ages (males 11 years, females 9 years). Height trajectories were visualized via LOESS smoothing. Between-group differences were tested using Kruskal-Wallis with Dunn post-hoc tests and bootstrap median differences. BMI transitions assessed whether weight-status normalization recovered height potential.

**Results:** At index ages, obese children were taller than normal-BMI peers (+5.1 cm at 11 years, peaking at +5.8 cm at 12 years for males and +4.0 cm at 9 years for females), while thin children were shorter (−3.0 to -4.2 cm). Differences narrowed progressively, becoming non-significant by 15 years in females (p = 0.885) and 17 years in males (p = 0.201). BMI normalization was associated with convergence toward normal-weight peers while persistent thinness showed no recovery. Results were consistent across ages, cities, and sensitivity analyses.

**Conclusions:** Pre-pubertal BMI shapes the tempo, not outcome, of pubertal height gain, supporting early, sex-specific nutritional intervention without compromising adult height.

**Key messages:**

- Pre-pubertal BMI shapes the tempo, not the final outcome, of pubertal height gain. Obese children are taller in childhood but this advantage resolves by mid-to-late adolescence, while thinness causes lasting shortfall unless BMI normalizes during puberty.
- This is the first large longitudinal Southeast Asian cohort to link BMI-transition patterns (not just cross-sectional BMI) to height recovery, filling a gap dominated by Western and East Asian studies.
- The study supports earlier, sex-specific nutritional intervention (before 11 years old for boys and 9 years old for girls) without compromising adult height.

## INTRODUCTION

Children’s linear growth and the onset of puberty are inextricably linked to nutritional status, and a growing body of evidence indicates that excess adiposity in early and middle childhood fundamentally alters the trajectory of height gain. Higher BMI in the pre-pubertal years is consistently associated with accelerated linear growth and advanced bone age during childhood, followed by an earlier onset of puberty, a shorter and attenuated pubertal growth spurt, and reduced height gain during adolescence ^1–4^. The biological mechanism involves multiple interconnected pathways: elevated leptin concentrations in children with excess adiposity stimulate the hypothalamic-pituitary-gonadal axis, advancing pubertal onset. Simultaneously, insulin-like growth factor 1 (IGF-1) and insulin signaling accelerate epiphyseal growth plate maturation, ultimately curtailing the duration of the pubertal growth window ^3,5^.

The consequences of undernutrition on linear growth are equally well established but mechanistically distinct. Children with thinness (BMI-for-age below -2 SD) and stunting (height-for-age below -2 SD) experience reduced growth velocity, delayed pubertal onset, and a prolonged growth window due to delayed growth plate closure, offering the theoretical possibility of compensatory or catch-up growth if nutritional deficits are addressed ^5,6^. In practice, evidence from multi-country LMIC cohorts demonstrates that faster linear growth and weight gain in childhood predict taller adult stature, although this is often accompanied by increased adiposity in adulthood ^7,8^. Body composition studies further indicate that skeletal muscle mass accretion rather than fat mass increment is the primary driver of successful catch-up growth in pre-pubertal children with short stature ^6^. These contrasting trajectories between undernutrition and overnutrition produce divergent patterns of height evolution across the childhood-to-adolescence continuum.

Beyond cross-sectional BMI category, the trajectory of BMI across childhood may carry independent prognostic significance for pubertal outcomes. A Taiwanese birth cohort study by Fan et al identified four BMI trajectory classes among children followed from age 6 to 18 years, finding that the chronically overweight/obese group (class 3) carried the highest risk of early pubertal maturation and the most divergent final height patterns by sex in that shorter stature in boys and taller stature in girls compared with healthy-weight peers ^9^. In Chinese children followed longitudinally from 2006 to 2016, Li et al demonstrated that earlier pubertal onset timing, smaller peak height velocity, and shorter pubertal spurt duration were each independently associated with lower final height and higher overweight and obesity risk in late adolescence, after controlling for baseline height via propensity score matching ^10^. Studies using QEPS growth modelling in the GrowUpGothenburg cohort have further developed pubertal-timing-aligned BMI references, confirming that individual variation in the age at pubertal onset substantially modifies the BMI-height relationship throughout adolescence ^11^.

Despite this growing body of evidence, the overwhelming majority of longitudinal studies characterizing the BMI–height trajectory relationship has been conducted in high-income European or North American populations or, more recently, in Chinese and Taiwanese cohorts ^1–4,9–12^. Evidence from Southeast Asia where children experience markedly different nutritional environments, genetic determinants of stature, pubertal timing, and trajectories of the nutrition transition compared with Western populations is very limited. Vietnamese children, in particular, face a uniquely compressed epidemiological transition, with stunting, thinness, and obesity coexisting across the same school-age population and the same calendar period ^13,14^. Whether the BMI–height trajectory relationship established in European cohorts generalizes to Vietnamese children, and whether the effect sizes and directional patterns differ by sex, age of BMI exposure, or city of residence, remains unknown.

Additional gaps persist. Most studies dichotomize children as overweight/obese versus normal weight, providing limited information on height trajectories in children with thinness, a category rarely considered despite its high LMIC prevalence. Few studies have examined whether children who change BMI category across the pubertal transition (e.g. obese children who normalize weight) recover height potential relative to persistently obese peers, a question with direct clinical and policy relevance for whether pre-pubertal weight management protects the pubertal growth trajectory.

This study retrospectively analyzed annual school health check data from children aged 18 months to 18 years attending a private school system in three major Vietnamese cities (Hanoi, Ho Chi Minh City, Haiphong) from 2018 to 2025. The aims were to: (1) characterize longitudinal height trajectories from childhood to late adolescence stratified by pre-pubertal BMI category, at sex-specific index ages (males 8 to 12 years, females 7 to 11 years); (2) quantify between-group height differences at each follow-up age; (3) examine whether children who normalize BMI across the pubertal transition recover height potential relative to persistently obese peers; and (4) assess consistency across three cities and both sexes.

## METHODS

### Study Design and Setting

This study analyzed annual school health check data of 249,745 child-year visits of 97,111 unique children aged from 18 months to 18 years attending a private school system in 3 major cities Hanoi, Hochiminh and Haiphong in Vietnam from 2018 to 2025. The 2021 data was not available due to COVID-19 restriction. Vinmec Ethical Committee (approval number 0231/2024/CN/HDDD VMEC) granted ethical approval and a waiver of individual informed consent as the study used de-identified routinely collected health data from school health examinations.

This study is reported in accordance with the STROBE statement (Strengthening the Reporting of Observational Studies in Epidemiology) for cohort studies. The completed STROBE checklist is provided as Supplementary Material.

### Study participants

A total of 97,111 unique children contributed 247,760 WHO-classified anthropometric observations to the full dataset. After applying the minimum three-visit criterion, the primary longitudinal analytic cohort comprised 40,887 children (177,545 person-visits). For subsequent modelling analyses, a further restriction was applied: children were required to have a valid WHO-referenced BMI classification at the designated pre-pubertal index age (age 11 years for males, age 9 years for females), yielding a male modelling sub-cohort of 6,410 boys (31,195 person-visits) and a female sub-cohort of 6,513 girls (30,729 person-visits). A sensitivity cohort restricted to children with four or more annual visits (n = 28,216) was used for all primary sensitivity analyses.

### WHO Reference-Based Anthropometric Classification

Prepubertal BMI status was classified using sex-specific WHO international reference standards applied by age group ^15,16^. BMI categories at baseline were defined as: thinness (BMI-for-age <□-2 SD), normal BMI (BMI-for-age ≥ -2□SD and ≤ +1□SD), overweight (BMI-for-age > +1□SD and ≤ +2□SD), and obesity (BMI-for-age > +2□SD). Normal BMI served as the reference category throughout. BMI category was treated as a time-varying covariate across annual observations and was classified independently at each visit. For trajectory analyses, the pre-pubertal index BMI category (a fixed, time-invariant exposure assigned from the observation recorded at the designated index age) was used as the primary exposure variable.

The primary outcome for all analyses was standing height (cm) measured at each annual examination. Height was modelled as a continuous variable in all regression and descriptive analyses.

### Exposure Definition: Pre-Pubertal Index BMI Category

#### Selection of Canonical Index Ages

The primary exposure was the WHO-referenced BMI category of each child at a designated pre-pubertal index age, defined as the age at which a representative snapshot of pre-pubertal nutritional status was captured before the onset of the pubertal growth acceleration. Based on established evidence on the timing of pubertal onset in Vietnamese children and in Asian populations more broadly, the canonical index ages were set at age 11 years for males and age 9 years for females. These ages precede mean pubertal onset by approximately 1 to 2 years in both sexes, ensuring that the index BMI reflects pre-pubertal body composition rather than pubertal body habitus.

To assess robustness to this choice, the full trajectory analysis was repeated for all adjacent pre-pubertal index ages: ages 8 through 12 years for males and ages 7 through 11 years for females.

#### BMI Trajectory Pairs

To characterize BMI status change across puberty and its association with height trajectories, children were additionally classified by their BMI transition pair which is defined as the combination of BMI category at an early pre-pubertal index age and a later post-pubertal or mid-pubertal age. The following transition windows were examined: males at ages 10→13 and 11→14 years, females at ages 8→11 and 9→12 years. Five trajectory pairs were defined: Obesity→Obesity, Obesity→Normal BMI, Normal BMI→Normal BMI, Thinness→Normal BMI, and Thinness→Thinness. Children whose BMI category at either the early or later time point was missing were excluded from the trajectory pair analysis for that window.

#### Statistical Analyses

All analyses were conducted in R version 4.5.1 ^17^. A two-sided p-value threshold of 0.05 was used for all hypothesis tests unless otherwise stated.

Simultaneous 95% confidence intervals for the four-category multinomial BMI distribution were computed using the Sison-Glaz method as implemented in the *DescTools* package (v0.99.60) ^18^. Population-level height trajectories by pre-pubertal BMI category were visualized using locally weighted scatterplot smoothing (LOESS) with 95% pointwise confidence interval shading.

Between-group height differences at each annual integer age (males: ages 8 to 18 years, females: ages 7 to 16 years) were evaluated using the non-parametric Kruskal-Wallis test. For ages at which the Kruskal-Wallis test was statistically significant, Dunn’s post-hoc pairwise comparisons were performed on all six pairs among the four BMI categories. Raw p-values from Dunn’s test were adjusted for multiple comparisons using the Benjamini-Hochberg false discovery rate (FDR) procedure ^19^. For each age and each comparison (thinness vs normal BMI, overweight vs normal BMI, obesity vs normal BMI), the observed difference in height medians and its 95% CI were estimated using the basic (percentile-based) bootstrap method with B = 999 replicates, as implemented in the *confintr* package (v1.0.2) ^20^.

The primary sensitivity analysis repeated all Kruskal-Wallis tests and trajectory visualizations restricting the cohort to children with four or more distinct annual visits (n = 28,216). This was performed to assess whether the inclusion of children with minimal longitudinal data (exactly three visits) influenced the observed trajectory patterns or convergence conclusions. Kruskal-Wallis statistics, median heights, and p-values at each milestone age were compared between the ≥3-visit and ≥4-visit analyses. Additional supportive analyses included: (1) city-stratified LOESS trajectory plots to assess geographic heterogeneity across Hanoi, Ho Chi Minh City, and Haiphong; (2) replication of the full trajectory analysis using all adjacent pre-pubertal index ages (ages 8 to 12 years for males, ages 7 to 11 years for females) to assess sensitivity to canonical index-age selection.

## RESULTS

### Study Population

Study population are summarized in **Table 1**.

**Table 1.**
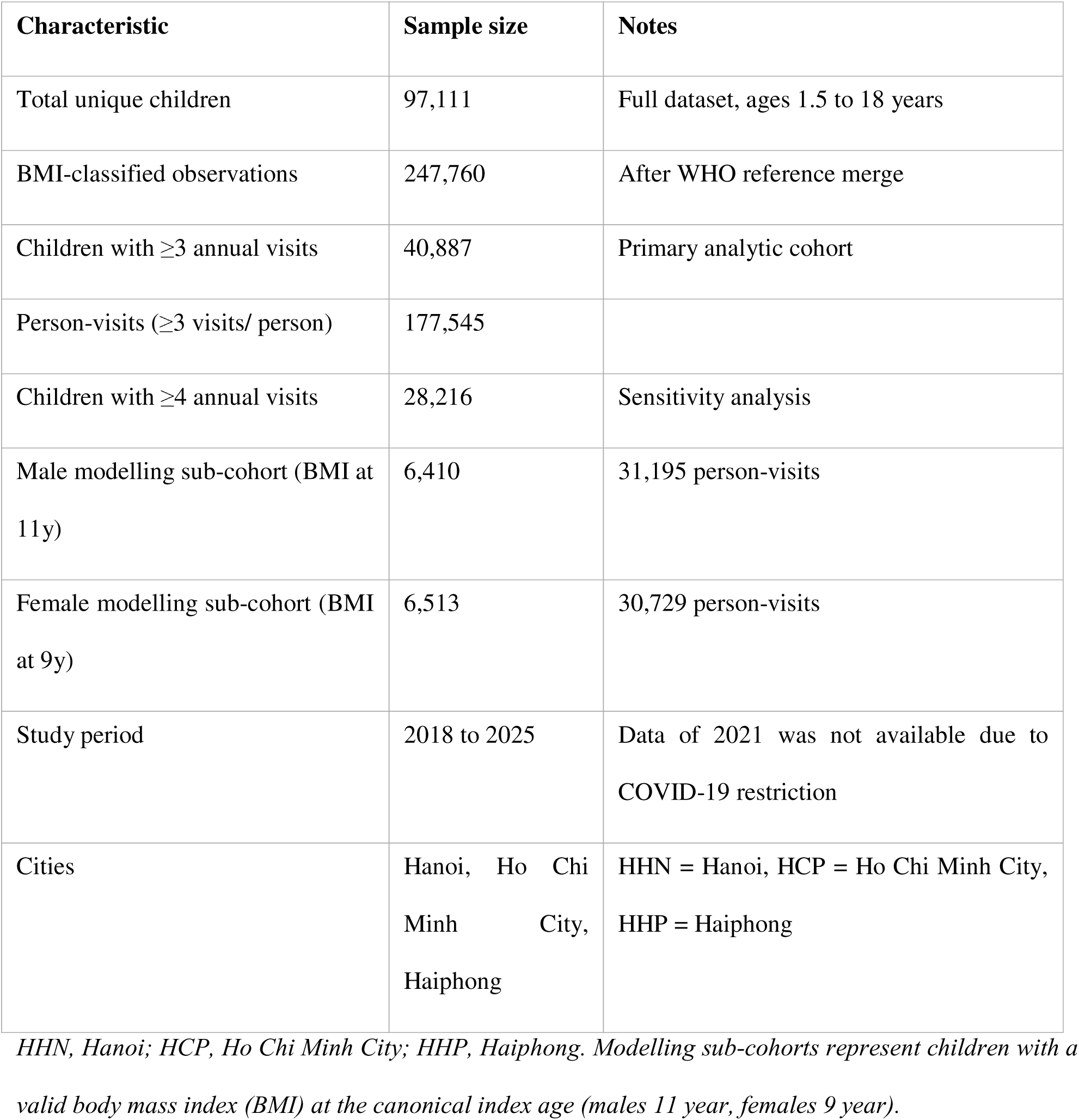
Summary of the longitudinal analytic cohort 2018 to 2025.

| Characteristic | Sample size | Notes |
| --- | --- | --- |
| Total unique children | 97,111 | Full dataset, ages 1.5 to 18 years |
| BMI-classified observations | 247,760 | After WHO reference merge |
| Children with $\geq 3$ annual visits | 40,887 | Primary analytic cohort |
| Person-visits ($\geq 3$ visits/ person) | 177,545 | |
| Children with $\geq 4$ annual visits | 28,216 | Sensitivity analysis |
| Male modelling sub-cohort (BMI at 11y) | 6,410 | 31,195 person-visits |
| Female modelling sub-cohort (BMI at 9y) | 6,513 | 30,729 person-visits |
| Study period | 2018 to 2025 | Data of 2021 was not available due to COVID-19 restriction |
| Cities | Hanoi, Ho Chi Minh City, Haiphong | HHN = Hanoi, HCP = Ho Chi Minh City, HHP = Haiphong |

*HHN, Hanoi; HCP, Ho Chi Minh City; HHP, Haiphong. Modelling sub-cohorts represent children with a valid body mass index (BMI) at the canonical index age (males 11 year, females 9 year).*

### Pre-Pubertal BMI Category Prevalence

**Table 2** presents the WHO-referenced BMI category distribution at the canonical pre-pubertal index ages and at adjacent ages for contextual comparison. At age 11 years in males, the prevalence of obesity was 29.5% (95% CI 28.1, 30.9) and overweight was 30.5% (29.1, 31.9), yielding a combined overweight–obesity burden of 60.0%. Normal BMI was present in 38.0% (36.6, 39.3) and thinness in 2.1% (0.7, 3.4). Notably, at age 8 years in males, normal BMI still predominated (47.9%), but had declined substantially by ages 9 to 11 years as obesity prevalence rose progressively (31.4% at age 9 years and 29.5% at age 11 years) (**Table S1**).

**Table 2.** Prevalence of WHO-referenced BMI categories and median height at canonical pre-pubertal index ages and contextual adjacent ages.

| Sex | Age (y) | BMI Category | N | BMI Category Prevalence % (95% CI) | Median height cm (95%CI) |
| --- | --- | --- | --- | --- | --- |
| Male | 8 | Thinness | 238 | 2.7 (1.6, 3.9) | 124.8 (124, 127) |
|  |  | Normal BMI | 4,159 | 47.9 (46.8, 49.1) | 128 (127.5, 128.5) |
|  |  | Overweight | 1,817 | 20.9 (19.8, 22.1) | 130 (130, 131) |
|  |  | Obesity | 2,460 | 28.4 (27.2, 29.5) | 131.8 (131.2, 132) |
|  | 11 | Thinness | 132 | 2.1 (0.7, 3.4) | 140.5 (139.7, 142) |
|  |  | Normal BMI | 2,434 | 38.0 (36.6, 39.3) | 143.7 (143.3, 144) |
|  |  | Overweight | 1,954 | 30.5 (29.1, 31.9) | 146 (146, 146.5) |
|  |  | Obesity | 1,890 | 29.5 (28.1, 30.9) | 148.8 (148.2, 149) |
|  | 16 | Thinness | 13 |  | 173 (167.4, 181) |
|  |  | Normal BMI | 291 |  | 171.7 (171, 172.5) |
|  |  | Overweight | 260 |  | 171 (170, 172) |
|  |  | Obesity | 232 |  | 172.5 (171.5, 173.4) |
|  | 17 | Thinness | 4 |  | 176.5 (-Inf, Inf) |
|  |  | Normal BMI | 82 |  | 173 (171, 175) |
|  |  | Overweight | 65 |  | 173.5 (172, 175) |
|  |  | Obesity | 56 |  | 174.2 (173, 176.5) |
| **Female** | 7 | Thinness | 246 | 3.8 (2.7, 4.9) | 119 (117.6, 120.5) |
|  |  | Normal BMI | 4,501 | 69.9 (68.8, 71.0) | 121 (121, 121) |
|  |  | Overweight | 1,153 | 17.9 (16.8, 19.0) | 123 (123, 124) |
|  |  | Obesity | 541 | 8.4 (7.3, 9.5) | 124 (123.2, 125) |
|  | 9 | Thinness | 243 | 3.7 (2.5, 5.0) | 130 (129.4, 131.7) |
|  |  | Normal BMI | 4,036 | 62.0 (60.8, 63.2) | 133 (132.5, 133) |
|  |  | Overweight | 1,604 | 24.6 (23.4, 25.9) | 135.5 (135, 136) |
|  |  | Obesity | 630 | 9.7 (8.5, 10.9) | 137 (136.2, 137.3) |
|  | 15 | Thinness | 6 |  | 158 (148.5, 168.5) |
|  |  | Normal BMI | 108 |  | 159.2 (158, 161.5) |
|  |  | Overweight | 50 |  | 159.5 (157, 161) |
|  |  | Obesity | 26 |  | 160.2 (156, 162) |
|  | 16 | Thinness | 6 |  | 162.2 (155, 167.5) |
|  |  | Normal BMI | 52 |  | 160 (158, 163) |
|  |  | Overweight | 25 |  | 160 (155, 162) |
|  |  | Obesity | 6 |  | 158.2 (154, 171) |

*Body mass index (BMI) categories defined by World Health Organization (WHO) 2006 weight-for-height references (age ≤5y) and WHO 2007 BMI-for-age references (age >5y). Estimates restricted to children with ≥3 annual visits and a valid BMI at the stated age. Sison-Glaz multinomial 95% confidence intervals (95%CI) for prevalence.*

In females at age 9 years, the BMI distribution was markedly different, with normal BMI constituting the majority at 62.0% (60.8, 63.2%) and combined overweight-obesity at 34.3% (overweight 24.6%, obesity 9.7%). At age 7 years, females had the highest normal BMI prevalence observed across any examined group (69.9%), with obesity at only 8.4%. The substantially lower obesity burden in pre-pubertal girls relative to boys of comparable age is a consistent feature of this urban Vietnamese school cohort and shapes the magnitude of height differences described below.

### Longitudinal Height Trajectories by Pre-Pubertal BMI Category

#### Population-Level LOESS Trajectories

**Figure 1** presents LOESS-smoothed height trajectories with 95% pointwise confidence intervals by pre-pubertal BMI category at the canonical index ages (BMI at 11 years for males, BMI at 9 years for females). Three consistent patterns emerged across both sexes. First, at the index age itself, height was ordered monotonically by BMI category: thinness < normal BMI < overweight < obesity. This gradient reflects the positive correlation between body mass and skeletal length throughout childhood. Second, through the early and mid-pubertal years, children with pre-pubertal overweight or obesity maintained or accelerated their height advantage, with LOESS trajectories remaining clearly separated and the 95% confidence bands non-overlapping between adjacent categories from approximately ages 8 to 14 years in males and 7 to 13 years in females. Third, in mid-to-late adolescence the trajectories converged markedly: by approximately age 15 to 16 years in males and age 13 to 14 years in females, the confidence intervals of the overweight and obesity groups overlapped those of the normal BMI group, and all four trajectories appeared to approach a common asymptote representing attained adult height.

**Figure 1.**
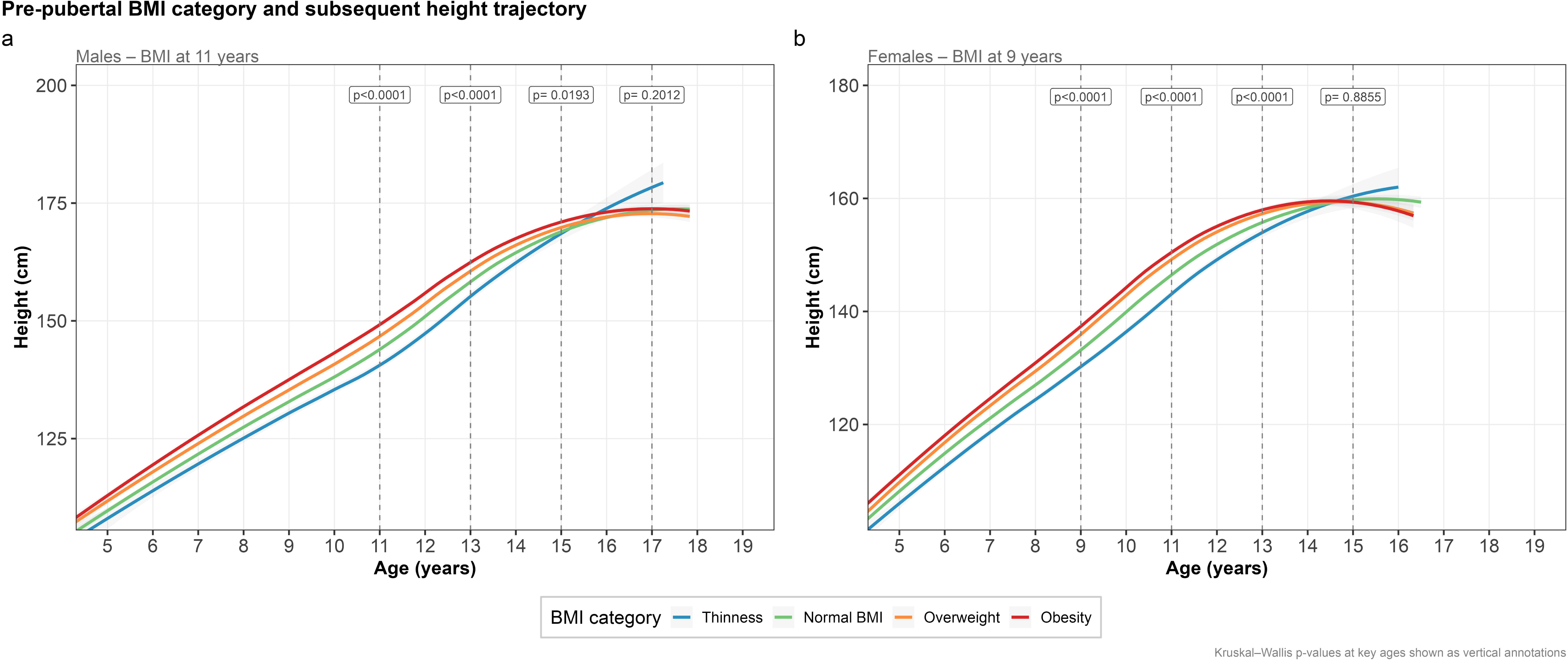
LOESS height trajectories annotated with Kruskal-Wallis p values at key milestone ages (dashed vertical lines). Panel a: Males (BMI at 11 years), with p values at ages 11, 13, 15, and 17 years. Panel b: Females (BMI at 9 years), with p values at ages 9, 11, 13, and 15 years. The sequential decline in KW statistics from highly significant (p < 0.0001) through borderline (p = 0.019 males; p < 0.0001 females at age 13) to non-significant (p = 0.2012 for males at 17 years, p = 0.8855 for females at 15 years) quantifies the trajectory convergence process.

The earlier convergence in females, visible as overlapping 95% CI bands from approximately age 13 years onward in **Figure 1b**, is consistent with the earlier timing of female puberty and the earlier completion of pubertal linear growth in girls relative to boys.

The consistency of these trajectory patterns across all five pre-pubertal index ages examined in each sex (ages 8 to 12 years for males, ages 7 to 11 years for females) is demonstrated in **Figure 2**. Regardless of which pre-pubertal year was chosen as the index, the three-phase pattern (early height advantage for higher BMI groups, pubertal amplification, late-adolescent convergence) was replicated in every panel, providing strong evidence against index-age selection bias. City-stratified analyses (**Figure 3**) confirmed that the pattern was broadly consistent across Hanoi, Ho Chi Minh City, and Haiphong, with the obesity group maintaining the tallest trajectory in each city during puberty.

**Figure 2.**
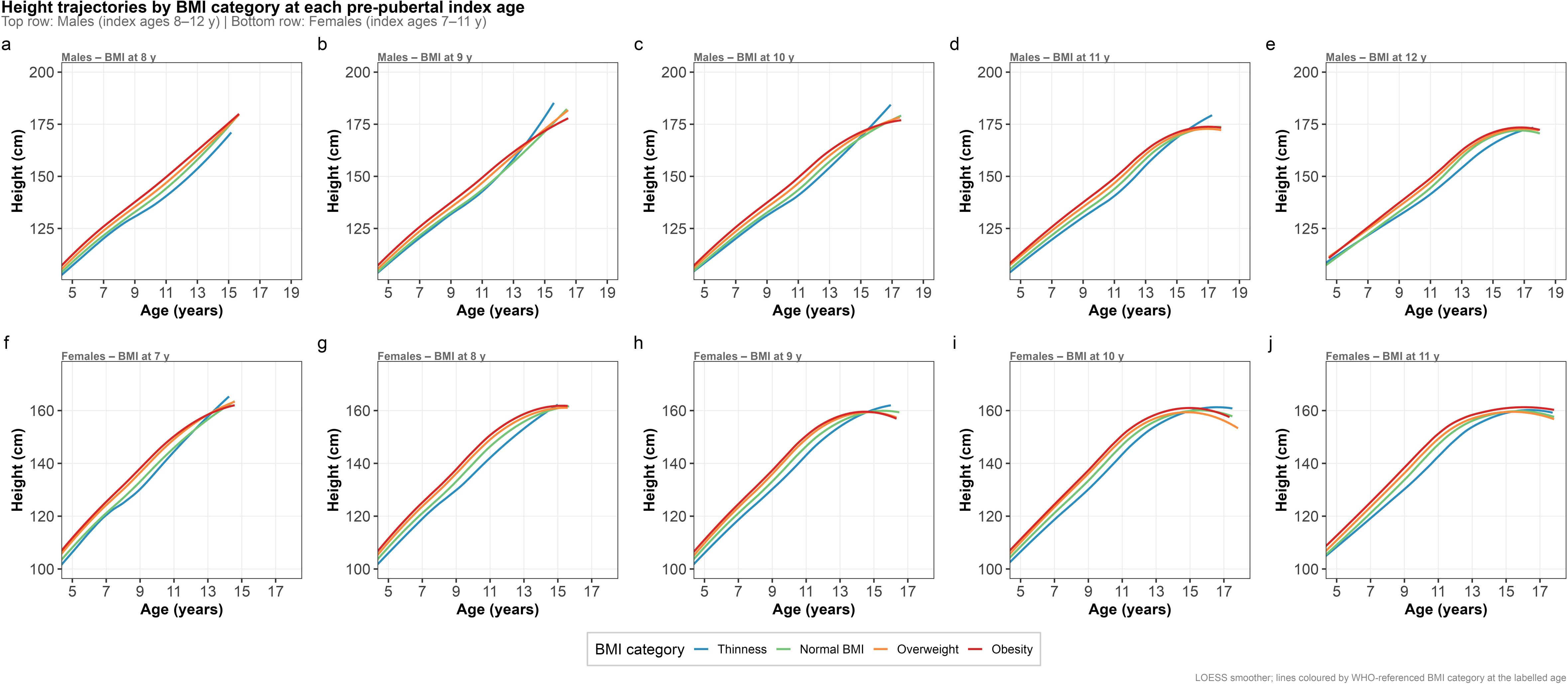
Height trajectories by WHO-referenced BMI category at all pre-pubertal index ages examined. Top row: males, index ages 8 to 12 years, panels a to e. Bottom row: females, index ages 7 to 11 years, panels f to j. LOESS smoothers without CI shading shown for visual clarity. The three-phase pattern (early height advantage, pubertal amplification, convergence) is reproduced consistently across all 10 panels, confirming that findings are not an artefact of canonical index-age selection.

**Figure 3.**
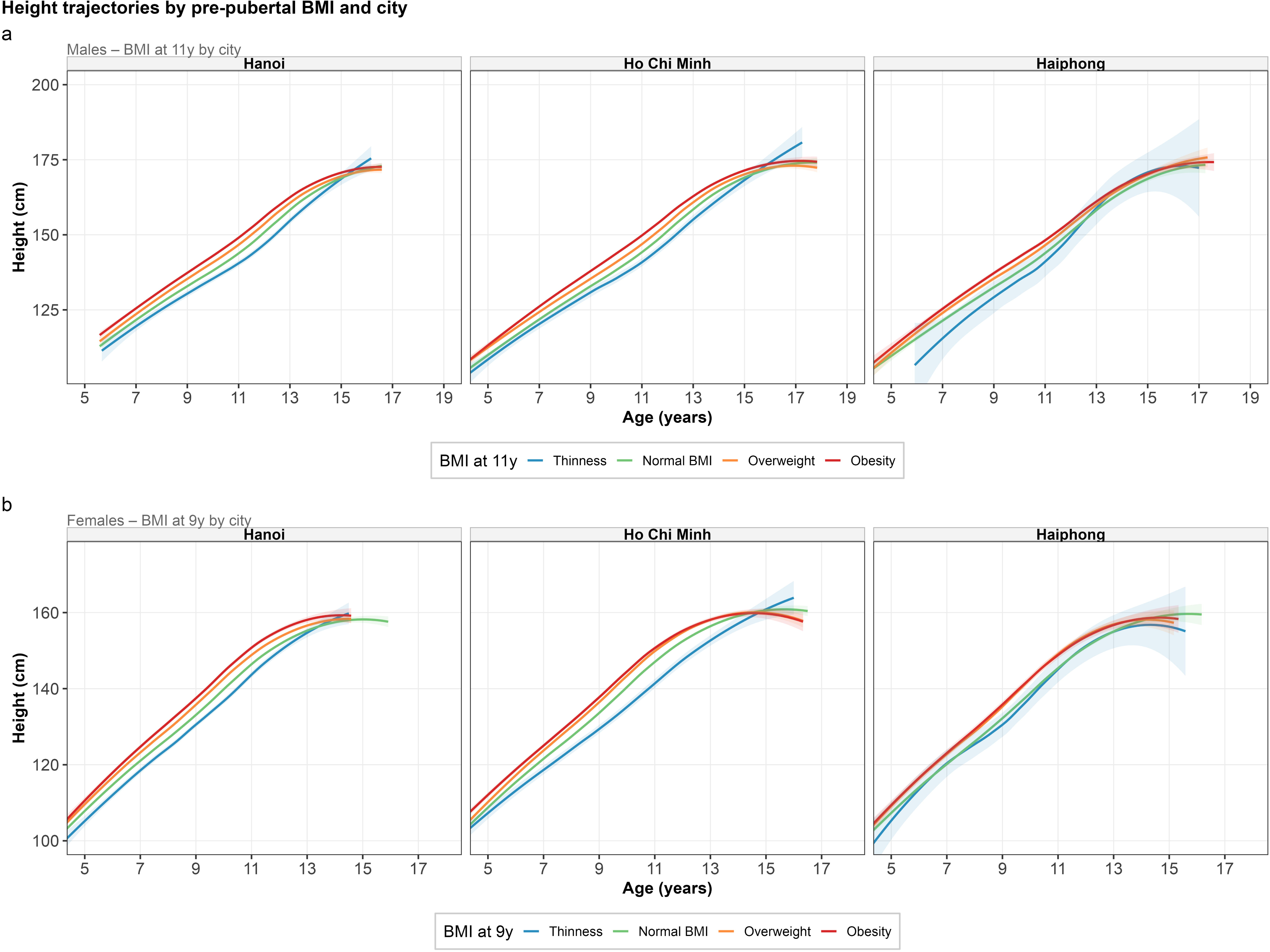
Height trajectories by pre-pubertal BMI category stratified by city of enrolment (Hanoi, Ho Chi Minh City, Haiphong). Panel a: Males, index BMI at age 11 years. Panel b: Females, index BMI at age 9 years. LOESS smoothers with 95% CI shading. The three-phase trajectory pattern (early height advantage for obesity ỏ overweight, pubertal amplification, late-adolescent convergence) is broadly consistent across all three cities, confirming that geographic variation within the VMEC does not drive the primary findings. Minor inter-city differences in absolute height levels are attributable to compositional and environmental factors.

#### Height Distributions at Milestone Ages

**Table 3** reports median heights for all BMI groups at each annual age from the pre-pubertal period through late adolescence and p-values from Kruskal-Wallis test (**Table S2**). **Figure 2** and **Figure 4** display the corresponding distributions and annotated trajectory plots.

**Figure 4.**
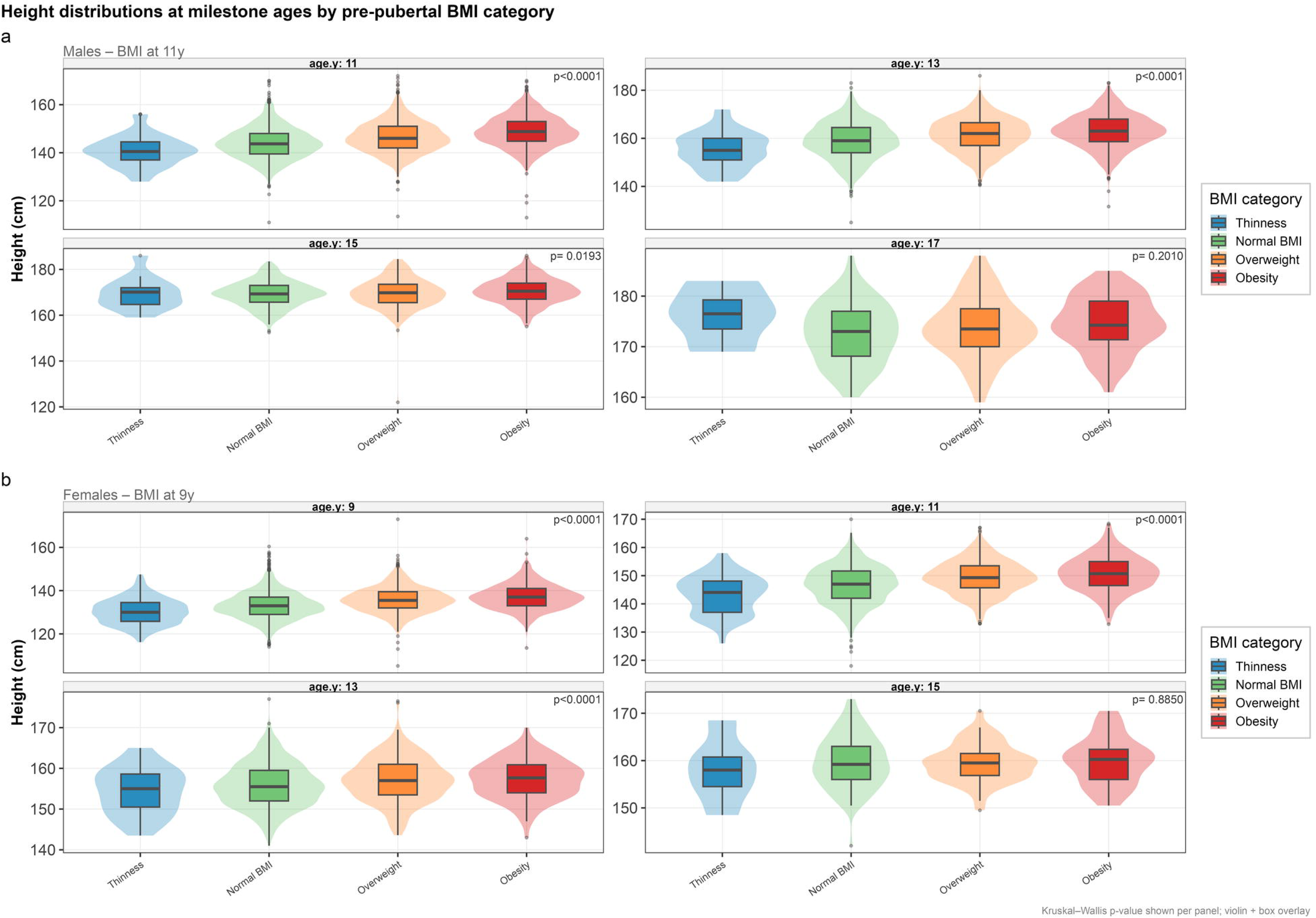
Height distributions at four milestone ages by pre-pubertal BMI category (violin plots with overlaid box-and-whisker plots). Panel a: Males (BMI at 11 years) at ages 11, 13, 15, and 17 years. Panel b: Females (BMI at 9 years) at ages 9, 11, 13, and 15 years. Kruskal–Wallis p values are annotated within each panel. The progressive compression of between-group differences and increasing distributional overlap from early to late adolescence is clearly visible. At age 17 years in males (p = 0.201) and age 15 years in females (p = 0.885), the violin envelopes are essentially superimposed.

**Table 3.** Median heights (cm) by pre-pubertal body mass index (BMI) category at each annual age.

| **Sex** | **Age**<br>**(y)** | **Thinness**<br>**Median**<br>**height cm** | **Normal**<br>**BMI**<br>**Median**<br>**height cm** | **Overweight**<br>**Median height**<br>**cm** | **Obesity**<br>**Median**<br>**height cm** | **p value** |
| --- | --- | --- | --- | --- | --- | --- |
| **Male** | 8 | 124.8 | 128.0 | 130.0 | 131.8 | <0.0001 |
|  | 9 | 131.0 | 133.0 | 135.0 | 137.0 | <0.0001 |
|  | 10 | 134.8 | 138.0 | 140.5 | 143.0 | <0.0001 |
|  | 11 | 140.5 | 143.7 | 146.0 | 148.8 | <0.0001 |
|  | 12 | 146.0 | 150.2 | 153.5 | 156.0 | <0.0001 |
|  | 13 | 155.0 | 159.0 | 162.0 | 163.0 | <0.0001 |
|  | 14 | 164.0 | 165.5 | 167.0 | 168.5 | <0.0001 |
|  | 15 | 170.1 | 169.3 | 169.8 | 170.5 | 0.019 |
|  | 16 | 173.0 | 171.7 | 171.0 | 172.5 | 0.152 |
|  | 17 | 176.5 | 173.0 | 173.5 | 174.2 | 0.201 |
| **Female** | 7 | 119.0 | 121.0 | 123.0 | 124.0 | <0.0001 |
|  | 8 | 123.4 | 126.5 | 129.5 | 130.4 | <0.0001 |
|  | 9 | 130.0 | 133.0 | 135.5 | 137.0 | <0.0001 |
|  | 10 | 136.0 | 140.0 | 143.0 | 144.0 | <0.0001 |
|  | 11 | 144.1 | 147.0 | 149.3 | 150.7 | <0.0001 |
|  | 12 | 150.5 | 152.3 | 154.0 | 155.0 | <0.0001 |
|  | 13 | 155.0 | 155.5 | 157.0 | 157.7 | <0.0001 |
|  | 14 | 158.0 | 157.5 | 159.0 | 159.0 | 0.009 |
|  | 15 | 158.0 | 159.2 | 159.5 | 160.2 | 0.885 |
|  | 16 | 162.2 | 160.0 | 160.0 | 158.2 | 0.428 |

In males (canonical index: BMI at 11 years), height differences were highly statistically significant at every age from 8 through 14 years (all p < 0.0001) (**Table 3**). At the index age of 11 years, median heights were 140.5 cm (thinness, n = 132), 143.7 cm (normal BMI, n = 2,434), 146.0 cm (overweight, n = 1,954), and 148.8 cm (obesity, n = 1,890) (p < 0.0001). The absolute spread between the thinness and obesity groups was 8.3 cm at age 11 years, widened further at age 12 years (10.0 cm: 146.0 vs 156.0 cm), then narrowed progressively. The violin-plot panels in **Figure 4** visualize this progressive compression: at ages 11 and 13 years, the four distributions are clearly separated with minimal overlap, whereas at age 15 years the distributions are largely superimposed. By age 17 years, the median heights of 176.5, 173.0, 173.5, and 174.2 cm for thinness, normal BMI, overweight, and obesity respectively (p = 0.201) were numerically indistinguishable, indicating complete statistical convergence of attained male height (**Figure 2**). In females (canonical index: BMI at 9 years), the pattern mirrored that of males but with accelerated timing. Significant differences were present from age 7 years (p < 0.0001) through age 14 years (p = 0.009). At the index age of 9 years, median heights were 130.0 cm (thinness), 133.0 cm (normal BMI), 135.5 cm (overweight), and 137.0 cm (obesity). The obesity-thinness spread at age 9 years was 7.0 cm, narrowing to 2.7 cm by age 13 years (155.0 vs 157.7 cm) and to 2.2 cm by age 14 years. By age 15 years, the median heights of 158.0, 159.2, 159.5, and 160.2 cm across the four groups showed near-complete numerical convergence (p = 0.885). At age 16 years, the point estimate for the obesity group median (158.2 cm) was numerically below that of the normal BMI group (160.0 cm), consistent with a marginal overshoot pattern in the smallest subgroup (n = 6), though this was not statistically significant (p = 0.428) (**Figure 1**, **Figure 3**, **Figure 4, Table S4**).

Dunn post-hoc pairwise comparisons with Benjamini-Hochberg correction confirmed that in males at age 13 years, all pairwise contrasts were significant (all BH-adjusted p < 0.002), including overweight vs obesity (p < 0.002). At age 16 years, all comparisons were non-significant, indicating that convergence was substantially complete by this age. In females, all pairwise contrasts at ages 9 and 11 years were significant (BH-adjusted p < 0.001), but by age 15 years no contrast was significant (**Table S3**).

### Magnitude of Height Differences

Bootstrap-derived median height differences relative to the normal BMI reference group quantify the trajectory patterns described above with age-specific precision (**Table 4** and **Figure 5, Table S6**).

**Figure 5.**
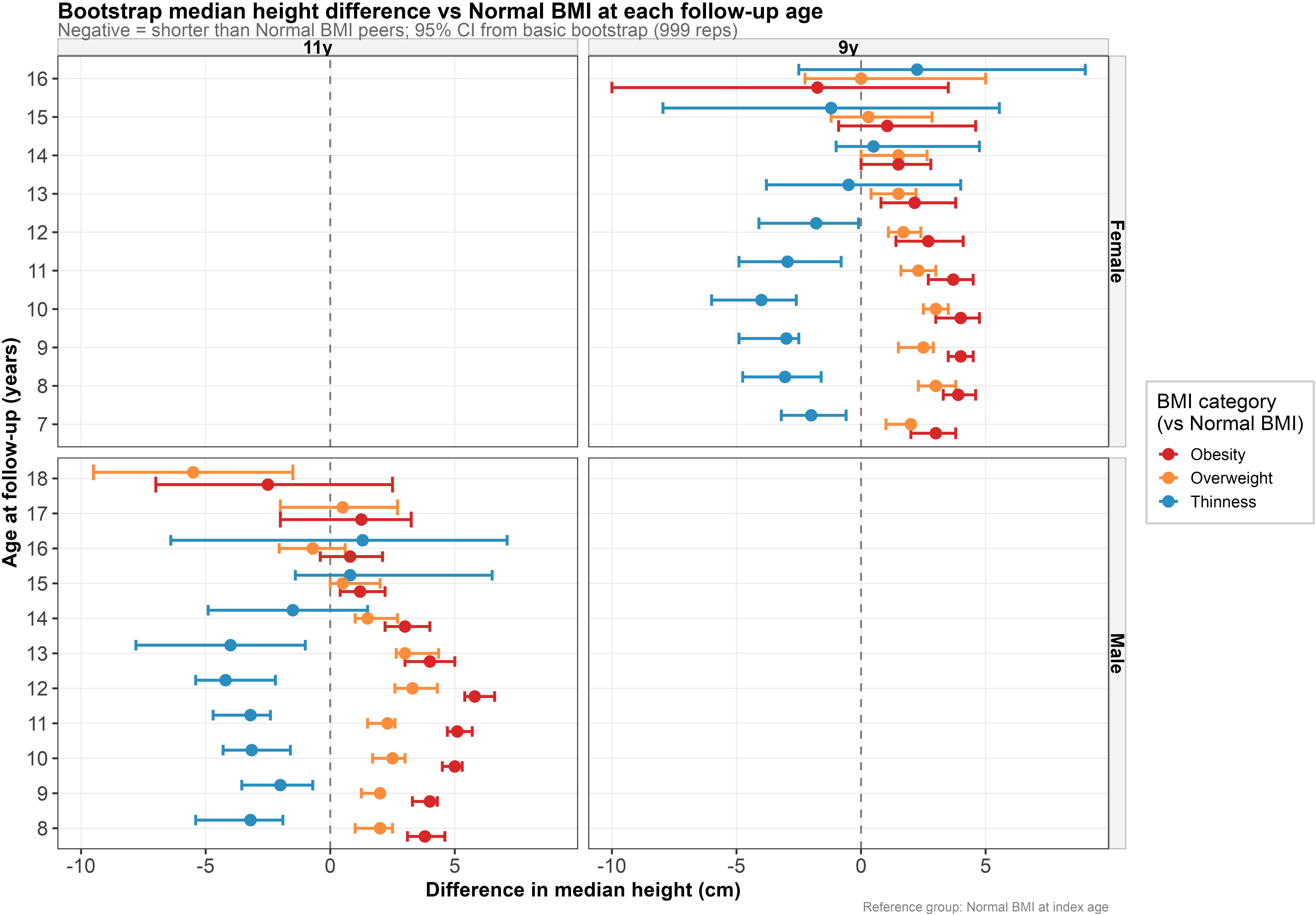
Forest-style plot of bootstrap median height differences (cm) versus the normal BMI reference group at each follow-up age (999 replicates, basic bootstrap; 95% CI). Left panel: Females, index BMI at age 9 years (ages 7 to 16y on y-axis). Right panel: Males, index BMI at age 11 years (ages 8 to 18y on y-axis). Color coding: Thinness (blue), Overweight (orange), Obesity (red). The vertical dashed line at zero indicates equivalence with normal BMI. The progressive convergence toward zero with increasing age is clearly visible for all comparisons in both sexes, with convergence occurring earlier in females. The obesity advantage and thinness deficit are approximately symmetric in magnitude at each age, consistent with both reflecting the same underlying relationship between pre-pubertal BMI and pubertal height tempo.

**Table 4.** Bootstrap median height differences (cm) relative to the normal BMI reference group at each follow-up age.

| Sex | Age (y) | Thinness vs Normal BMI Median difference cm (95% CI) | Overweight vs Normal BMI Median difference cm (95% CI) | Obesity vs Normal BMI Median difference cm (95% CI) |
| --- | --- | --- | --- | --- |
| Male | 8 | -3.2 (-5.4, -1.9) | +2.0 (+1.0, +2.5) | +3.8 (+3.1, +4.6) |
|  | 9 | -2.0 (-3.9, -0.7) | +2.0 (+1.3, +2.0) | +4.0 (+3.3, +4.3) |
|  | 10 | -3.2 (-4.3, -1.7) | +2.5 (+1.8, +3.0) | +5.0 (+4.5, +5.3) |
|  | 11 | -3.2 (-4.7, -2.4) | +2.3 (+1.6, +2.6) | +5.1 (+4.7, +5.8) |
|  | 12 | -4.2 (-4.9, -2.2) | +3.3 (+2.6, +4.4) | +5.8 (+5.4, +6.6) |
|  | 13 | -4.0 (-7.8, -1.2) | +3.0 (+2.8, +4.4) | +4.0 (+3.1, +5.0) |
|  | 14 | -1.5 (-4.9, +3.0) | +1.5 (+1.0, +2.7) | +3.0 (+2.2, +4.0) |
|  | 15 | +0.8 (-1.4, +6.6) | +0.5 (0.0, +1.8) | +1.2 (+0.4, +2.2) |
|  | 16 | +1.3 (-5.9, +7.1) | -0.7 (-2.1, +0.6) | +0.8 (-0.4, +2.1) |
|  | 17 |  | +0.5 (-2.0, +3.0) | +1.3 (-2.3, +3.0) |
| Female | 7 | -2.0 (-3.2, -0.6) | +2.0 (+1.0, +2.1) | +3.0 (+2.0, +3.8) |
|  | 8 | -3.1 (-4.8, -1.6) | +3.0 (+2.3, +3.8) | +3.9 (+3.2, +4.7) |
|  | 9 | -3.0 (-4.7, -2.5) | +2.5 (+1.5, +2.9) | +4.0 (+3.5, +4.5) |
|  | 10 | -4.0 (-6.0, -2.7) | +3.0 (+2.5, +3.5) | +4.0 (+3.0, +4.6) |
|  | 11 | -3.0 (-4.9, -0.9) | +2.3 (+1.6, +3.0) | +3.7 (+2.9, +4.5) |
|  | 12 | -1.8 (-4.1, -0.1) | +1.7 (+1.0, +2.4) | +2.7 (+1.4, +4.1) |
|  | 13 | -0.5 (-4.0, +3.9) | +1.5 (+0.4, +2.4) | +2.2 (+0.8, +3.8) |
|  | 14 | +0.5 (-1.0, +4.3) | +1.5 (0.0, +2.5) | +1.5 (0.0, +2.7) |
|  | 15 | -1.2 (-7.7, +5.5) | +0.3 (-1.2, +3.1) | +1.1 (-0.9, +4.6) |

*Negative values indicate shorter stature relative to normal BMI peers; positive values indicate taller stature. Confidence intervals crossing zero indicate non-significant differences. This denotes insufficient sample size ( $n < 5$ ) for estimation. All estimates are unadjusted nonparametric median differences. Bootstrap with 999 replicates, basic bootstrap method; 95% CI. Males: index BMI at age 11 years. Females: index BMI at age 9 years.*

For thinness males, the deficit was present from age 8 years (−3.2 cm, -5.4 to -1.9) and remained stable through age 11 years (−3.2 cm, -4.7 to -2.4), widening to 4.2 cm (−4.9, -2.2) at age 12 and 4.0 cm (−7.8, -1.2) at age 13. By age 14 the deficit narrowed to -1.5 cm (−4.9, +3.0), non-significant. At ages 15 to 17 years, point estimates turned positive (+0.8 to +1.3 cm) with wide non-significant intervals, indicating catch-up by mid-adolescence.

For overweight males, a 2.3 cm advantage at age 11 (+1.6, +2.6) rose to 3.3 cm (+2.6, +4.4) at age 12, then diminished progressively: 3.0 cm (+2.8, +4.4) at 13, 1.5 cm (+1.0, +2.7) at 14, and 0.5 cm (0.0, +1.8) at 15, with the lower bound reaching zero. By age 16 the estimate turned negative (−0.7 cm), non-significant, indicating complete convergence.

For obesity males, the height advantage was largest overall: 5.1 cm (+4.7, +5.8) at age 11, roughly half an expected annual increment, peaking at 5.8 cm (+5.4, +6.6) at age 12. This contracted to 4.0 cm (+3.1, +5.0) at 13, 3.0 cm (+2.2, +4.0) at 14, and 1.2 cm (+0.4, +2.2) at 15. By age 17 the interval spanned zero (+1.3 cm, -2.3, +3.0), indicating equivalent attained adult height. **Figure 5** (right panel) shows this as a consistent leftward shift of obesity estimates with age.

In females, parallel patterns emerged roughly 2 years earlier. The thinness deficit peaked at age 10 (−4.0 cm, -6.0 to -2.7), became non-significant by age 13 (−0.5 cm, -4.0 to +3.9), and reversed slightly at ages 14–16 with zero-spanning intervals. The obesity advantage peaked at age 9 (+4.0 cm, +3.5 to +4.5), declining to +2.7 cm (+1.4, +4.1) at age 12 and +1.1 cm (−0.9, +4.6) at age 15, crossing zero. This leftward shift appears earlier in **Figure 5** (left panel) than in males, consistent with earlier pubertal maturation. The symmetric obesity-thinness crossing pattern in **Figure 5** confirms that the pubertal height advantage of obesity is transient, mirrored by the transient deficit of thinness.

### Height Trajectories by BMI Category Transition Across Puberty

**Figure 6** presents height trajectories stratified by BMI status at two time points spanning puberty: males at ages 10→13 and 11→14 years (panels a, c), females at ages 8→11 and 9→12 years (panels b, d). Five trajectory pairs are shown: Obesity→Obesity, Obesity→Normal BMI, Normal BMI→Normal BMI, Thinness→Normal BMI, and Thinness→Thinness. Children with persistent obesity were tallest during early to mid-puberty in both sexes and transition windows, but by late adolescence (∼17 years in males, ∼15 in females) converged toward the Normal BMI→Normal BMI group. In some panels, the Obesity→Normal BMI group ended at comparable or slightly greater final heights, consistent with earlier weight normalization being associated with a modestly more favorable late-adolescent trajectory.

**Figure 6.**
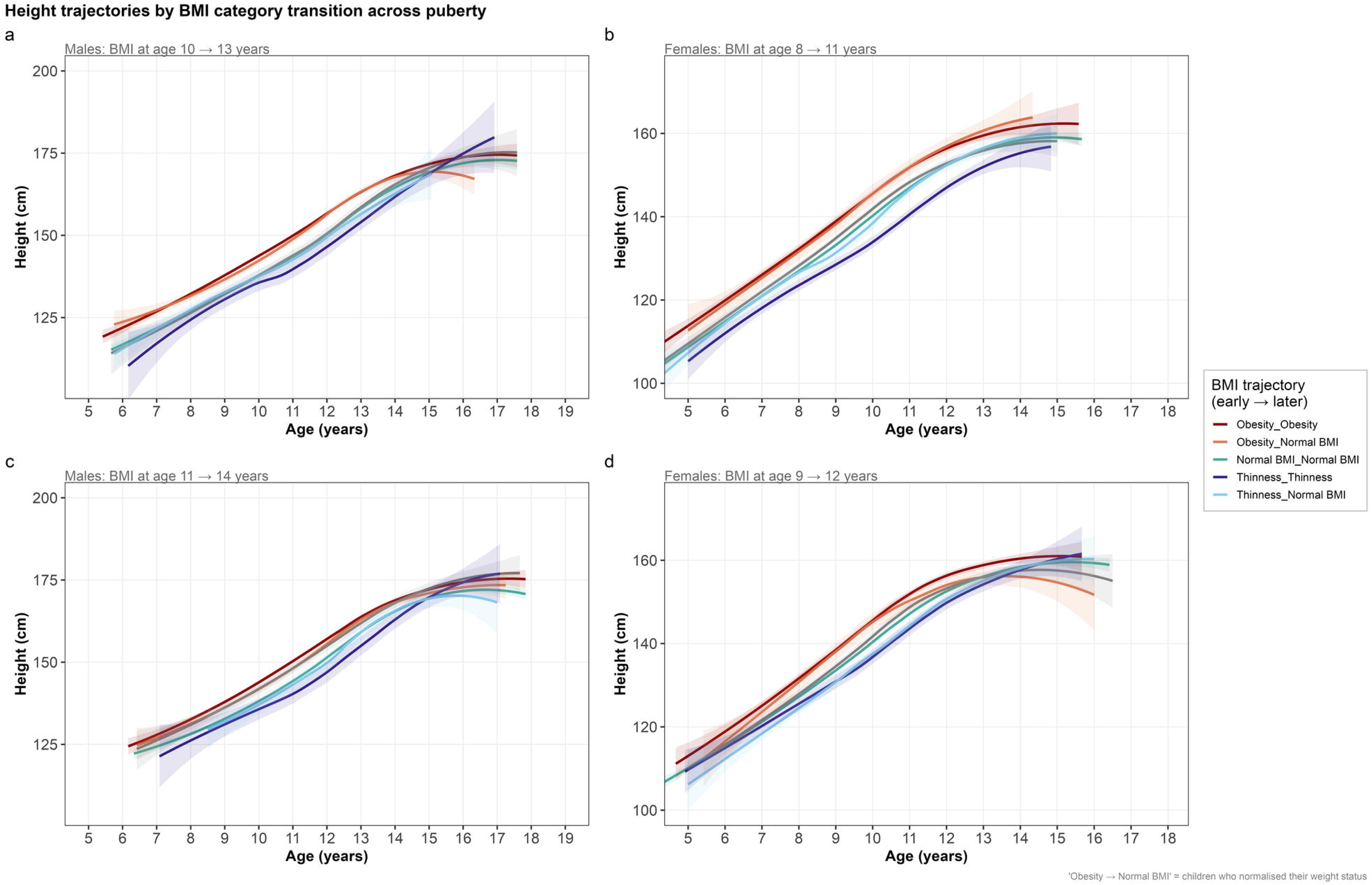
Height trajectories by BMI category transition pair across puberty. Panel a: Males, BMI transition age 10→13 years. Panel b: Females, BMI transition age 8→11 years. Panel c: Males, BMI transition age 11→14 years. Panel d: Females, BMI transition age 9→12 years. Trajectory pairs are color-coded as shown in the legend. LOESS smoothers with 95% CI shading. ’Obesity→Normal BMI’ represents children who normalized weight status across the transition window. ’Thinness→Thinness’ represents children with persistent undernutrition. The visible convergence of all trajectories in late adolescence (except for persistent thinness) is consistent with the nonparametric and mixed-model analyses.

Children with persistent thinness showed consistently the shortest trajectories and no compensatory catch-up. By contrast, Thinness→Normal BMI trajectories converged toward the Normal BMI→Normal BMI group by late adolescence, suggesting partial height catch-up with recovery from thinness. This pattern is consistent across both transition windows in each sex.

The Normal BMI→Normal BMI group followed intermediate trajectories, reaching final heights consistent with Kruskal-Wallis convergence values (males ∼173 to 174 cm, females ∼158 to 160 cm). Overall, **Figure 6** reinforces that late-adolescent height outcomes converge across most trajectory patterns, though persistent thinness may carry the greatest risk of suboptimal attained height, while persistent obesity confers taller pre-pubertal rather than final stature.

### Sensitivity Analyses

Restricting the analytic cohort to children with four or more annual visits (n = 28,216, approximately 69% of the primary cohort) produced findings that were virtually identical to the primary analysis (**Table S5**). In males, the convergence pattern was precisely replicated: p = 0.019 at age 15 years and p = 0.201 at age 17 years which are identical to the main analysis values. In females, significance at ages 13 years (p < 0.0001) and 14 years (p = 0.009) was confirmed, and non-significance at ages 15 years (p = 0.885) and 16 years (p = 0.428) was maintained without change. Median heights by BMI group were within 0.2 cm of main-analysis values at all ages. These findings confirm that neither selective attrition of children with fewer visits nor differences in follow-up intensity materially influenced the reported trajectory patterns or statistical conclusions.

## DISCUSSION

To our knowledge, this is the first longitudinal multi-city study characterizing the relationship between pre-pubertal BMI status and height trajectories across childhood to late adolescence in a Southeast Asian population. Using annual school health data from 40,887 Vietnamese children (177,545 person-visits from Hanoi, Ho Chi Minh City, Haiphong from 2018 to 2025), we found a consistent three-phase pattern in both sexes: childhood height advantage among overweight or obese children, narrowing through mid-puberty, and near-complete convergence by late adolescence (∼15 years in females, ∼17 in males). Pre-pubertal thinness showed the inverse, incompletely-resolved pattern. Findings were reproducible across index ages, cities, and a sensitivity cohort.

### Pre-Pubertal Obesity and the Transient Height Advantage

Higher pre-pubertal BMI is consistently associated with accelerated linear growth and advanced bone age, followed by earlier pubertal onset, a shorter growth spurt, and reduced adolescent height gain ^1–4^. Elevated leptin in children with excess adiposity stimulates the hypothalamic-pituitary-gonadal axis, advancing pubertal onset, while IGF-1 and insulin signaling accelerate epiphyseal maturation, curtailing the pubertal growth window ^4,5^.

In our cohort, the obesity-group height advantage was largest at the index age (+5.1 cm at 11 years in males, +4.0 cm at 9 years in females), peaking roughly a year later (+5.8 cm at 12 years in males) and this is consistent with these children already entering pubertal acceleration before peers. Convergence, confirmed by LOESS visualization, Kruskal-Wallis testing, and bootstrap median differences, is the central, most clinically relevant finding. The mechanism mirrors the early advantage: earlier puberty drives earlier epiphyseal closure, shortening the height-gain window so early acceleration is not sustained into late adolescence.

### Pre-Pubertal Thinness and BMI Transitions

Children with thinness showed the inverse trajectory, with deficits widening to 4.2 cm (males, 12 years) and 4.0 cm (females, 10 years) before narrowing. Confidence intervals crossed zero roughly 3 years earlier than for the obesity advantage, but point estimates remained slightly negative, leaving catch-up uncertain rather than confirmed. Mechanistically, undernutrition suppresses GH-IGF-1 signaling and delays HPG-axis activation via reduced leptin and kisspeptin ^5^, producing delayed but theoretically extended pubertal growth. This is consistent with multi-country LMIC evidence that nutritional rehabilitation before or during early puberty supports partial catch-up growth ^7,8^.

Our BMI-transition-pair analysis supports this: children who normalized from thinness to normal BMI converged toward normal-weight peers by late adolescence, while persistent thinness showed no equivalent recovery. This is a clinically actionable distinction not captured by cross-sectional BMI category alone. Chun et al., in a Korean cohort with pre-pubertal short stature, similarly found lean-mass (not fat-mass) accretion drove height catch-up ^6^, suggesting that body composition change, not BMI normalization per se, may be the more specific predictor of recovery.

The same transition-pair analysis showed that children who normalized from obesity to normal BMI reached final heights comparable to, or marginally exceeding, those of persistently obese peers. This is consistent with Putri et al. ^12^ and reinforcing that BMI normalization, in either direction, does not compromise and may modestly favor attained adult height. Together, the findings suggest that the pre-pubertal and early pubertal years constitute the critical window during which nutritional status, whether excess or deficit, can still be modified without lasting consequence for final height.

### Clinical Implications

These findings support two complementary clinical messages. For obesity, weight management initiated before puberty (before age 11 years in boys and 9 years in girls in this population) is not harmful to linear growth and should not be deferred over fears of stunting final height. This is consistent with Putri et al.’s treatment-cohort evidence ^12^ and with the demonstration here that BMI normalization does not reduce attained height relative to persistently obese peers. Given that combined overweight-obesity reached 60.0% in 11-year-old boys, the pre-pubertal years represent a narrow window in which intervention can moderate the early pubertal height advantage while improving cardiometabolic risk, without compromising adult stature.

For thinness, nutritional support should begin before the pubertal window closes, since catch-up was demonstrated only in children who normalized BMI during puberty, not those remaining thin throughout. Height-for-age monitoring alone is insufficient: it may falsely reassure clinicians about overweight children (who appear tall throughout childhood) while masking reduced growth potential in thin children whose height remains nominally normal despite falling short of genetic potential. Concurrent height- and BMI-for-age surveillance, attentive to the pre-pubertal detection windows identified here (9 years in girls, 11 years in boys), is recommended for school health systems in similarly transitioning Southeast Asian and other LMIC populations.

### Strengths and Limitations

Strengths include the longitudinal data with large sample size, three-city geographic diversity, a robust ≥4-visit sensitivity analysis confirming all findings, and convergent evidence from three methods replicated across five pre-pubertal index ages in each sex.

Several limitations should be noted. First, the absence of direct pubertal staging data (Tanner stage, age at menarche) within the school health records means the proposed pubertal-timing mechanism is inferred from trajectory shape rather than directly confirmed. Second, the private-school cohort is not representative of the general Vietnamese child population, limiting generalizability and constraining precision for the smaller thinness subgroups, particularly at older ages (n = 4 at age 17y in males). Third, late-adolescent attrition as children graduate or drop out from the school network reduces statistical power for the oldest age comparisons. Fourth, no parental height data were available to adjust for genetic height potential, though He and Karlberg found that the childhood BMI–pubertal height gain association persisted after such adjustment in their Swedish cohort ^2^, offering some reassurance that the relationship reported here is not purely genetic in origin. Fifth, BMI also does not distinguish fat from lean mass, so some misclassification of the overweight category cannot be excluded, although this is less likely to affect the obesity category given its very high prevalence in this cohort. Finally, formal growth modelling with prediction of individual trajectories will be presented in a separate paper. The present descriptive findings provide the empirical foundation for that subsequent work.

## Conclusions

In this large Vietnamese school-based cohort (the first such study from Southeast Asia spanning childhood to late adolescence), pre-pubertal BMI category strongly shaped pubertal height trajectories but not attained adult height. Overweight and obese children were taller during puberty but converged to heights indistinguishable from normal-weight peers by age 15 (females) and 17 (males). Children with pre-pubertal thinness remained shortest throughout, with recovery seen only when BMI normalized during puberty. Given the very high overweight-obesity prevalence (60.0% in 11-year-old boys) and reassurance that BMI normalization does not compromise adult height, pre-pubertal nutritional intervention for both obesity and thinness should be prioritized in school health policy.

### Data Sharing Statement

Aggregated summary statistics are provided as supplementary tables. R codes are available from the corresponding author upon reasonable request. Individual patient-level data cannot be shared due to applicable privacy regulations and the terms of the institutional ethics approval.

## Contributors’ statement

Nhan T. Ho did conceptualization, data curation, formal analysis, investigation, methodology, project administration, resources, software, supervision, validation, visualization, writing original draft, and writing review & editing. Michelle Hermiston, Quang N. Nguyen, Lam N. Phung, and Cuong T. Do did supervision, writing review & editing. Quynh T. Nguyen, Minh A. Nguyen, Quyet V. Nguyen, Anh Q. Dao, Chi T. L. Tran, and Nhan T. Ho did data curation, resources, validation. An N. Pham did data curation, resources, supervision, validation, and writing review & editing. All authors read and approved the manuscript.

## Conflict of intertest

On behalf of all authors, the corresponding author states that there is no conflict of interest

## Consent statement

The requirement for individual informed consent was waived by the Vinmec Ethical Committee (approval number 0231/2024/CN/HDDD VMEC) because this study used de-identified routinely collected health data from school health examinations.

## Use of Artificial Intelligence

All scientific content, analyses, and interpretations are the original work of the authors. The authors used AI-assisted tool (Paperpal) for language editing and grammar checking during manuscript preparation.

## REFERENCES

1. Holmgren, A. et al. Pubertal height gain is inversely related to peak BMI in childhood. Pediatr. Res. 81, 448–454 (2017).

2. He, Q. & Karlberg, J. BMI in Childhood and Its Association with Height Gain, Timing of Puberty, and Final Height. Pediatr. Res. 49, 244–251 (2001).

3. Shalitin, S. & Gat-Yablonski, G. Associations of Obesity with Linear Growth and Puberty. Horm. Res. Paediatr. 95, 120–136 (2021).

4. Aris, I., et al. Association of BMI with Linear Growth and Pubertal Development. Obesity 27, (2019).

5. Soliman, A. et al. Impact of BMI on childhood growth, pubertal timing, and bone maturation: A comprehensive review and clinical implications. World Journal of Advanced Research and Reviews (2024) doi:10.30574/wjarr.2024.23.3.2854.

6. Chun, D., Kim, S. J., Suh, J. & Kim, J. Body Composition Changes and Catch-up Growth in Pre-pubertal Children with Short Stature: A Longitudinal Retrospective Cross-sectional Cohort Study. J. Clin. Res. Pediatr. Endocrinol. 17, 458–467 (2025).

7. Adair, L. et al. Associations of linear growth and relative weight gain during early life with adult health and human capital in countries of low and middle income: findings from five birth cohort studies. Lancet 382, 525–534 (2013).

8. Poveda, N. et al. Growth patterns in childhood and adolescence and adult body composition: a pooled analysis of birth cohort studies from five low and middle-income countries (COHORTS collaboration). BMJ Open 13, (2023).

9. Fan, H.-Y., Lee, Y., Hsieh, R., Yang, C. & Chen, Y. Body mass index growth trajectories, early pubertal maturation, and short stature. Pediatr. Res. 88, 117–124 (2019).

10. Chen, L. et al. Association between height growth patterns in puberty and stature in late adolescence: A longitudinal analysis in chinese children and adolescents from 2006 to 2016. Front. Endocrinol. (Lausanne). 13, (2022).

11. Albertsson-Wikland, K., Niklasson, A., Gelander, L., Holmgren, A. & Nierop, A. Novel type of references for BMI aligned for onset of puberty – using the QEPS growth model. BMC Pediatr. 22, (2022).

12. Putri, R. R., Danielsson, P., Marcus, C. & Hagman, E. Height and Growth Velocity in Children and Adolescents Undergoing Obesity Treatment: A Prospective Cohort Study. J. Clin. Endocrinol. Metab. 109, e314–e320 (2023).

13. Ho, N. T. et al. Overweight & obesity epidemic, temporal trends and regional disparities in physical growth of Vietnamese children. Sci. Rep. (2026).

14. Mai, T. M. T. et al. The double burden of malnutrition in Vietnamese school-aged children and adolescents: a rapid shift over a decade in Ho Chi Minh City. Eur. J. Clin. Nutr. 74, 1448–1456 (2020).

15. De Onis, M. et al. Development of a WHO growth reference for school-aged children and adolescents. Bull. World Health Organ. 85, (2007).

16. WHO Multicentre Growth Reference Study Group. WHO Child Growth Standards: Length/Height-for-Age, Weight-for-Age, Weight-for-Length, Weight-for-Height and Body Mass Index-for-Age: Methods and Development. (World Health Organization, Geneva, 2006).

17. R Core Team. R: A Language and Environment for Statistical Computing. R Foundation for Statistical Computing, Vienna, Austria Preprint at 10.1017/CBO9781107415324.004 (2016).

18. Signorell, A. DescTools: Tools for Descriptive Statistics. (2025). doi:10.32614/CRAN.package.DescTools.

19. Benjamini, Y. & Hochberg, Y. Controlling the false discovery rate: a practical and powerful approach to multiple testing. Journal of the Royal Statistical Society Series B 57, 289–300 (1995).

20. Mayer, M. Confintr: Confidence Intervals. (2023). doi:10.32614/CRAN.package.confintr.

