## Supplementary Materials for "Effect of Pre-Pubertal Body Mass Index Status on Longitudinal Height Trajectory in Vietnamese Children: 2018-2025 School Health Analysis"

Nhan Thi Ho et al.

**List of supplementary Tables**

**Table S1.** Prevalence of body mass index (BMI) category at index age.

**Table S2**. Comparison of height between body mass index (BMI) categories at index age (boy=11, girl= 9 years) at various age.

**Table S3**. Post-hoc Dunn test.

**Table S4**. Median height difference between body mass index (BMI) categories at index age.

**Table S5**. Sensitivity analysis on children with ≥4 visits.

**Table S6**. Median height and 95%CI by body mass index (BMI) category, sex and age.

**Supplementary Tables**

**Table S1.** Prevalence of body mass index (BMI) category at index age.

| **Sex** | **Age** | **BMI Category** | **N** | **Prevalence % (95%CI)** |
| --- | --- | --- | --- | --- |
| Male | 8 | Thinness | 238 | 2.7 (1.6, 3.9) |
| Male | 8 | Normal BMI | 4159 | 47.9 (46.8, 49.1) |
| Male | 8 | Overweight | 1817 | 20.9 (19.8, 22.1) |
| Male | 8 | Obesity | 2460 | 28.4 (27.2, 29.5) |
| Male | 9 | Thinness | 164 | 2.3 (1.0, 3.5) |
| Male | 9 | Normal BMI | 3017 | 41.8 (40.5, 43.0) |
| Male | 9 | Overweight | 1777 | 24.6 (23.3, 25.9) |
| Male | 9 | Obesity | 2266 | 31.4 (30.1, 32.6) |
| Male | 10 | Thinness | 161 | 2.1 (0.9, 3.4) |
| Male | 10 | Normal BMI | 2860 | 37.5 (36.2, 38.7) |
| Male | 10 | Overweight | 2174 | 28.5 (27.2, 29.7) |
| Male | 10 | Obesity | 2438 | 31.9 (30.7, 33.2) |
| Male | 11 | Thinness | 132 | 2.1 (0.7, 3.4) |
| Male | 11 | Normal BMI | 2434 | 38.0 (36.6, 39.3) |
| Male | 11 | Overweight | 1954 | 30.5 (29.1, 31.9) |
| Male | 11 | Obesity | 1890 | 29.5 (28.1, 30.9) |
| Male | 12 | Thinness | 165 | 2.5 (1.2, 3.8) |
| Male | 12 | Normal BMI | 2814 | 42.3 (41.0, 43.7) |
| Male | 12 | Overweight | 2135 | 32.1 (30.8, 33.5) |
| Male | 12 | Obesity | 1532 | 23.1 (21.7, 24.4) |
| Female | 7 | Thinness | 246 | 3.8 (2.7, 4.9) |
| Female | 7 | Normal BMI | 4501 | 69.9 (68.8, 71.0) |
| Female | 7 | Overweight | 1153 | 17.9 (16.8, 19.0) |
| Female | 7 | Obesity | 541 | 8.4 (7.3, 9.5) |
| Female | 8 | Thinness | 264 | 3.4 (2.3, 4.4) |
| Female | 8 | Normal BMI | 5155 | 65.7 (64.6, 66.8) |
| Female | 8 | Overweight | 1677 | 21.4 (20.3, 22.4) |
| Female | 8 | Obesity | 753 | 9.6 (8.5, 10.7) |
| Female | 9 | Thinness | 243 | 3.7 (2.5, 5.0) |
| Female | 9 | Normal BMI | 4036 | 62.0 (60.8, 63.2) |
| Female | 9 | Overweight | 1604 | 24.6 (23.4, 25.9) |
| Female | 9 | Obesity | 630 | 9.7 (8.5, 10.9) |
| Female | 10 | Thinness | 305 | 4.3 (3.1, 5.4) |
| Female | 10 | Normal BMI | 4418 | 61.8 (60.7, 63.0) |
| Female | 10 | Overweight | 1808 | 25.3 (24.1, 26.5) |
| Female | 10 | Obesity | 615 | 8.6 (7.5, 9.8) |
| Female | 11 | Thinness | 247 | 4.0 (2.8, 5.2) |
| Female | 11 | Normal BMI | 3997 | 64.6 (63.4, 65.8) |
| Female | 11 | Overweight | 1523 | 24.6 (23.4, 25.8) |
| Female | 11 | Obesity | 420 | 6.8 (5.6, 8.0) |

95%CI: 95% confidence interval.

**Table S2**. Comparison of height between body mass index (BMI) categories at index age (boy=11, girl= 9 years) at various age.

| **Sex** | **Index BMI age** | **Age (y)** | **Median height (cm) by group** | **KW p** |
| --- | --- | --- | --- | --- |
| Male | 11y | 8 | Thinness:124.8(n=58); Normal BMI:128(n=1017); Overweight:130(n=812); Obesity:131.8(n=819) | <0.001 |
| Male | 11y | 9 | Thinness:131(n=69); Normal BMI:133(n=1228); Overweight:135(n=991); Obesity:137(n=951) | <0.001 |
| Male | 11y | 10 | Thinness:134.8(n=90); Normal BMI:138(n=1587); Overweight:140.5(n=1248); Obesity:143(n=1217) | <0.001 |
| Male | 11y | 11 | Thinness:140.5(n=132); Normal BMI:143.7(n=2434); Overweight:146(n=1954); Obesity:148.8(n=1890) | <0.001 |
| Male | 11y | 12 | Thinness:146(n=89); Normal BMI:150.2(n=1552); Overweight:153.5(n=1331); Obesity:156(n=1261) | <0.001 |
| Male | 11y | 13 | Thinness:155(n=61); Normal BMI:159(n=1186); Overweight:162(n=1026); Obesity:163(n=978) | <0.001 |
| Male | 11y | 14 | Thinness:164(n=35); Normal BMI:165.5(n=875); Overweight:167(n=747); Obesity:168.5(n=697) | <0.001 |
| Male | 11y | 15 | Thinness:170.1(n=13); Normal BMI:169.3(n=365); Overweight:169.8(n=325); Obesity:170.5(n=299) | 0.0193 |
| Male | 11y | 16 | Thinness:173(n=13); Normal BMI:171.7(n=291); Overweight:171(n=260); Obesity:172.5(n=232) | 0.1517 |
| Male | 11y | 17 | Thinness:176.5(n=4); Normal BMI:173(n=82); Overweight:173.5(n=65); Obesity:174.2(n=56) | 0.2012 |
| Male | 11y | 18 | Normal BMI:176.5(n=19); Overweight:171(n=16); Obesity:174(n=13) | 0.0412 |
| Female | 9y | 7 | Thinness:119(n=133); Normal BMI:121(n=2144); Overweight:123(n=851); Obesity:124(n=342) | <0.001 |
| Female | 9y | 8 | Thinness:123.4(n=160); Normal BMI:126.5(n=2736); Overweight:129.5(n=1097); Obesity:130.4(n=427) | <0.001 |
| Female | 9y | 9 | Thinness:130(n=243); Normal BMI:133(n=4036); Overweight:135.5(n=1604); Obesity:137(n=630) | <0.001 |
| Female | 9y | 10 | Thinness:136(n=166); Normal BMI:140(n=2601); Overweight:143(n=1055); Obesity:144(n=418) | <0.001 |
| Female | 9y | 11 | Thinness:144.1(n=100); Normal BMI:147(n=1839); Overweight:149.3(n=779); Obesity:150.7(n=306) | <0.001 |
| Female | 9y | 12 | Thinness:150.5(n=81); Normal BMI:152.3(n=1381); Overweight:154(n=574); Obesity:155(n=230) | <0.001 |
| Female | 9y | 13 | Thinness:155(n=31); Normal BMI:155.5(n=631); Overweight:157(n=300); Obesity:157.7(n=110) | 3E-05 |
| Female | 9y | 14 | Thinness:158(n=30); Normal BMI:157.5(n=554); Overweight:159(n=256); Obesity:159(n=90) | 0.0091 |
| Female | 9y | 15 | Thinness:158(n=6); Normal BMI:159.2(n=108); Overweight:159.5(n=50); Obesity:160.2(n=26) | 0.8855 |
| Female | 9y | 16 | Thinness:162.2(n=6); Normal BMI:160(n=52); Overweight:160(n=25); Obesity:158.2(n=6) | 0.4276 |

KW p: p-values from Kruskal-Wallis test.

**Table S3**. Post-hoc Dunn test.

| **Age (year)** | **Measure** | **Group 1** | **Group 2** | **N 1** | **N 2** | **p-value** | **Adjusted p-value #** | **Sex** |
| --- | --- | --- | --- | --- | --- | --- | --- | --- |
| 11 | height | Thinness | Normal BMI | 132 | 2434 | 5.1224E-07 | 5.1224E-07 | Male |
| 11 | height | Thinness | Overweight | 132 | 1954 | 6.1313E-21 | 7.3576E-21 | Male |
| 11 | height | Thinness | Obesity | 132 | 1890 | 3.1241E-40 | 9.3724E-40 | Male |
| 11 | height | Normal BMI | Overweight | 2434 | 1954 | 9.4534E-39 | 1.8907E-38 | Male |
| 11 | height | Normal BMI | Obesity | 2434 | 1890 | 5.672E-131 | 3.403E-130 | Male |
| 11 | height | Overweight | Obesity | 1954 | 1890 | 1.398E-27 | 2.0969E-27 | Male |
| 13 | height | Thinness | Normal BMI | 61 | 1186 | 0.00047849 | 0.00047849 | Male |
| 13 | height | Thinness | Overweight | 61 | 1026 | 7.1149E-09 | 1.0672E-08 | Male |
| 13 | height | Thinness | Obesity | 61 | 978 | 1.2547E-13 | 3.7642E-13 | Male |
| 13 | height | Normal BMI | Overweight | 1186 | 1026 | 9.548E-13 | 1.9096E-12 | Male |
| 13 | height | Normal BMI | Obesity | 1186 | 978 | 2.5665E-33 | 1.5399E-32 | Male |
| 13 | height | Overweight | Obesity | 1026 | 978 | 1.4697E-06 | 1.7637E-06 | Male |
| 15 | height | Thinness | Normal BMI | 13 | 365 | 0.97630071 | 0.97630071 | Male |
| 15 | height | Thinness | Overweight | 13 | 325 | 0.79851419 | 0.95821703 | Male |
| 15 | height | Thinness | Obesity | 13 | 299 | 0.38298699 | 0.604096 | Male |
| 15 | height | Normal BMI | Overweight | 365 | 325 | 0.40273067 | 0.604096 | Male |
| 15 | height | Normal BMI | Obesity | 365 | 299 | 0.00220418 | 0.01322508 | Male |
| 15 | height | Overweight | Obesity | 325 | 299 | 0.02900573 | 0.0870172 | Male |
| 17 | height | Thinness | Normal BMI | 4 | 82 | 0.22462708 | 0.44188951 | Male |
| 17 | height | Thinness | Overweight | 4 | 65 | 0.36824126 | 0.44188951 | Male |
| 17 | height | Thinness | Obesity | 4 | 56 | 0.57768509 | 0.57768509 | Male |
| 17 | height | Normal BMI | Overweight | 82 | 65 | 0.3405841 | 0.44188951 | Male |
| 17 | height | Normal BMI | Obesity | 82 | 56 | 0.05428362 | 0.3257017 | Male |
| 17 | height | Overweight | Obesity | 65 | 56 | 0.33612699 | 0.44188951 | Male |
| 9 | height | Thinness | Normal BMI | 243 | 4036 | 2.0412E-09 | 2.4495E-09 | Female |
| 9 | height | Thinness | Overweight | 243 | 1604 | 4.0039E-35 | 6.0059E-35 | Female |
| 9 | height | Thinness | Obesity | 243 | 630 | 1.9845E-45 | 3.969E-45 | Female |
| 9 | height | Normal BMI | Overweight | 4036 | 1604 | 1.1166E-53 | 3.3497E-53 | Female |
| 9 | height | Normal BMI | Obesity | 4036 | 630 | 1.683E-55 | 1.0098E-54 | Female |
| 9 | height | Overweight | Obesity | 1604 | 630 | 3.9345E-06 | 3.9345E-06 | Female |
| 11 | height | Thinness | Normal BMI | 100 | 1839 | 4.0681E-06 | 4.8817E-06 | Female |
| 11 | height | Thinness | Overweight | 100 | 779 | 6.6026E-16 | 9.9039E-16 | Female |
| 11 | height | Thinness | Obesity | 100 | 306 | 8.6711E-20 | 2.6013E-19 | Female |
| 11 | height | Normal BMI | Overweight | 1839 | 779 | 2.1832E-19 | 4.3663E-19 | Female |
| 11 | height | Normal BMI | Obesity | 1839 | 306 | 1.137E-20 | 6.8222E-20 | Female |
| 11 | height | Overweight | Obesity | 779 | 306 | 0.00471224 | 0.00471224 | Female |
| 13 | height | Thinness | Normal BMI | 31 | 631 | 0.23697633 | 0.28437159 | Female |
| 13 | height | Thinness | Overweight | 31 | 300 | 0.00986319 | 0.01479478 | Female |
| 13 | height | Thinness | Obesity | 31 | 110 | 0.00810849 | 0.01479478 | Female |
| 13 | height | Normal BMI | Overweight | 631 | 300 | 0.00012306 | 0.00073833 | Female |
| 13 | height | Normal BMI | Obesity | 631 | 110 | 0.00190358 | 0.00571073 | Female |
| 13 | height | Overweight | Obesity | 300 | 110 | 0.64397788 | 0.64397788 | Female |
| 15 | height | Thinness | Normal BMI | 6 | 108 | 0.4454421 | 0.8882001 | Female |
| 15 | height | Thinness | Overweight | 6 | 50 | 0.50163982 | 0.8882001 | Female |
| 15 | height | Thinness | Obesity | 6 | 26 | 0.43866789 | 0.8882001 | Female |
| 15 | height | Normal BMI | Overweight | 108 | 50 | 0.8619496 | 0.8882001 | Female |
| 15 | height | Normal BMI | Obesity | 108 | 26 | 0.8882001 | 0.8882001 | Female |
| 15 | height | Overweight | Obesity | 50 | 26 | 0.8025611 | 0.8882001 | Female |

\# Multiple testing adjusted p-value using the Benjamini-Hochberg false discovery rate (FDR) procedure.

**Table S4**. Median height difference between body mass index (BMI) categories at index age.

| **Age (y)** | **Comparison (vs Normal BMI)** | **Median difference (cm) 95%CI** | **Sex** | **Index age (year)** |
| --- | --- | --- | --- | --- |
| 8 | Thinness | -3.2 (-5.4, -1.9) | Male | 11 |
| 8 | Overweight | 2 (1, 2.5) | Male | 11 |
| 8 | Obesity | 3.8 (3.1, 4.6) | Male | 11 |
| 9 | Thinness | -2 (-3.55, -0.7) | Male | 11 |
| 9 | Overweight | 2 (1.25, 2) | Male | 11 |
| 9 | Obesity | 4 (3.3, 4.3) | Male | 11 |
| 10 | Thinness | -3.15 (-4.3, -1.6) | Male | 11 |
| 10 | Overweight | 2.5 (1.7, 3) | Male | 11 |
| 10 | Obesity | 5 (4.5, 5.3) | Male | 11 |
| 11 | Thinness | -3.2 (-4.7, -2.4) | Male | 11 |
| 11 | Overweight | 2.3 (1.5, 2.6) | Male | 11 |
| 11 | Obesity | 5.1 (4.7, 5.7) | Male | 11 |
| 12 | Thinness | -4.2 (-5.4, -2.2) | Male | 11 |
| 12 | Overweight | 3.3 (2.6, 4.3) | Male | 11 |
| 12 | Obesity | 5.8 (5.4, 6.6) | Male | 11 |
| 13 | Thinness | -4 (-7.8, -1) | Male | 11 |
| 13 | Overweight | 3 (2.65, 4.35) | Male | 11 |
| 13 | Obesity | 4 (3, 5) | Male | 11 |
| 14 | Thinness | -1.5 (-4.9, 1.5) | Male | 11 |
| 14 | Overweight | 1.5 (1, 2.7) | Male | 11 |
| 14 | Obesity | 3 (2.2, 4) | Male | 11 |
| 15 | Thinness | 0.8 (-1.4, 6.5) | Male | 11 |
| 15 | Overweight | 0.5 (0, 2) | Male | 11 |
| 15 | Obesity | 1.2 (0.4, 2.2) | Male | 11 |
| 16 | Thinness | 1.3 (-6.4, 7.1) | Male | 11 |
| 16 | Overweight | -0.7 (-2.05, 0.6) | Male | 11 |
| 16 | Obesity | 0.8 (-0.4, 2.1) | Male | 11 |
| 17 | Overweight | 0.5 (-2, 2.7) | Male | 11 |
| 17 | Obesity | 1.25 (-2, 3.25) | Male | 11 |
| 18 | Overweight | -5.5 (-9.5, -1.5) | Male | 11 |
| 18 | Obesity | -2.5 (-7, 2.5) | Male | 11 |
| 7 | Thinness | -2 (-3.2, -0.6) | Female | 9 |
| 7 | Overweight | 2 (1, 2) | Female | 9 |
| 7 | Obesity | 3 (2, 3.8) | Female | 9 |
| 8 | Thinness | -3.05 (-4.75, -1.6) | Female | 9 |
| 8 | Overweight | 3 (2.3, 3.8) | Female | 9 |
| 8 | Obesity | 3.9 (3.3, 4.6) | Female | 9 |
| 9 | Thinness | -3 (-4.9, -2.5) | Female | 9 |
| 9 | Overweight | 2.5 (1.5, 2.9) | Female | 9 |
| 9 | Obesity | 4 (3.5, 4.5) | Female | 9 |
| 10 | Thinness | -4 (-6, -2.6) | Female | 9 |
| 10 | Overweight | 3 (2.5, 3.5) | Female | 9 |
| 10 | Obesity | 4 (3, 4.75) | Female | 9 |
| 11 | Thinness | -2.95 (-4.9, -0.8) | Female | 9 |
| 11 | Overweight | 2.3 (1.6, 3) | Female | 9 |
| 11 | Obesity | 3.7 (2.7, 4.5) | Female | 9 |
| 12 | Thinness | -1.8 (-4.1, -0.1) | Female | 9 |
| 12 | Overweight | 1.7 (1.1, 2.4) | Female | 9 |
| 12 | Obesity | 2.7 (1.4, 4.1) | Female | 9 |
| 13 | Thinness | -0.5 (-3.8, 4) | Female | 9 |
| 13 | Overweight | 1.5 (0.4, 2.2) | Female | 9 |
| 13 | Obesity | 2.15 (0.8, 3.8) | Female | 9 |
| 14 | Thinness | 0.5 (-1, 4.75) | Female | 9 |
| 14 | Overweight | 1.5 (0, 2.65) | Female | 9 |
| 14 | Obesity | 1.5 (0, 2.8) | Female | 9 |
| 15 | Thinness | -1.2 (-7.95, 5.55) | Female | 9 |
| 15 | Overweight | 0.3 (-1.2, 2.85) | Female | 9 |
| 15 | Obesity | 1.05 (-0.9, 4.6) | Female | 9 |
| 16 | Thinness | 2.25 (-2.5, 9) | Female | 9 |
| 16 | Overweight | 0 (-2.25, 5) | Female | 9 |
| 16 | Obesity | -1.75 (-10, 3.5) | Female | 9 |

**Table S5**. Sensitivity analysis on children with ≥4 visits.

| **Sex** | **Index BMI age** | **Age (y)** | **Median height (cm) by group** | **KW p- value #** |
| --- | --- | --- | --- | --- |
| Male | 11y (≥4 vis) | 8 | Thinness:124.6(n=55); Normal BMI:128(n=937); Overweight:130(n=769); Obesity:132(n=760) | 0 |
| Male | 11y (≥4 vis) | 9 | Thinness:130.5(n=60); Normal BMI:133(n=1121); Overweight:135(n=931); Obesity:137(n=873) | 0 |
| Male | 11y (≥4 vis) | 10 | Thinness:134.4(n=80); Normal BMI:138(n=1404); Overweight:140.5(n=1145); Obesity:143(n=1102) | 0 |
| Male | 11y (≥4 vis) | 11 | Thinness:140.2(n=111); Normal BMI:143.6(n=2063); Overweight:146(n=1744); Obesity:149(n=1629) | 0 |
| Male | 11y (≥4 vis) | 12 | Thinness:145.8(n=78); Normal BMI:150(n=1369); Overweight:153.5(n=1230); Obesity:156(n=1131) | 0 |
| Male | 11y (≥4 vis) | 13 | Thinness:154.3(n=55); Normal BMI:159(n=1061); Overweight:162(n=944); Obesity:163(n=871) | 0 |
| Male | 11y (≥4 vis) | 14 | Thinness:163.8(n=34); Normal BMI:166(n=818); Overweight:167(n=714); Obesity:168.5(n=660) | 0 |
| Male | 11y (≥4 vis) | 15 | Thinness:170.1(n=13); Normal BMI:169.3(n=365); Overweight:169.8(n=324); Obesity:170.5(n=299) | 0.019 |
| Male | 11y (≥4 vis) | 16 | Thinness:173(n=13); Normal BMI:171.7(n=291); Overweight:171(n=260); Obesity:172.5(n=232) | 0.1517 |
| Male | 11y (≥4 vis) | 17 | Thinness:176.5(n=4); Normal BMI:173(n=82); Overweight:173.5(n=65); Obesity:174.2(n=56) | 0.2012 |
| Male | 11y (≥4 vis) | 18 | Normal BMI:176.5(n=19); Overweight:171(n=16); Obesity:174(n=13) | 0.0412 |
| Female | 9y (≥4 vis) | 7 | Thinness:119(n=117); Normal BMI:121(n=1908); Overweight:123(n=781); Obesity:124(n=308) | 0 |
| Female | 9y (≥4 vis) | 8 | Thinness:123.3(n=127); Normal BMI:126.6(n=2345); Overweight:129.6(n=980); Obesity:130.5(n=375) | 0 |
| Female | 9y (≥4 vis) | 9 | Thinness:130(n=193); Normal BMI:133(n=3420); Overweight:135.6(n=1402); Obesity:137(n=543) | 0 |
| Female | 9y (≥4 vis) | 10 | Thinness:135(n=139); Normal BMI:140(n=2291); Overweight:143(n=951); Obesity:144(n=371) | 0 |
| Female | 9y (≥4 vis) | 11 | Thinness:144.9(n=90); Normal BMI:147(n=1704); Overweight:149.3(n=723); Obesity:150.6(n=282) | 0 |
| Female | 9y (≥4 vis) | 12 | Thinness:151(n=76); Normal BMI:152.5(n=1333); Overweight:154(n=555); Obesity:154.8(n=220) | 0 |
| Female | 9y (≥4 vis) | 13 | Thinness:155(n=31); Normal BMI:155.5(n=626); Overweight:157(n=300); Obesity:157.7(n=110) | 2E-05 |
| Female | 9y (≥4 vis) | 14 | Thinness:158(n=30); Normal BMI:157.5(n=554); Overweight:159(n=256); Obesity:159(n=90) | 0.0091 |
| Female | 9y (≥4 vis) | 15 | Thinness:158(n=6); Normal BMI:159.2(n=108); Overweight:159.5(n=50); Obesity:160.2(n=26) | 0.8855 |
| Female | 9y (≥4 vis) | 16 | Thinness:162.2(n=6); Normal BMI:160(n=52); Overweight:160(n=25); Obesity:158.2(n=6) | 0.4276 |

#p-values from Kruskal Wallis test.

**Table S6**. Median height and 95%CI by body mass index (BMI) category, sex and age.

| **Sex** | **Age (y)** | **BMI category** | **N** | **Median height cm (95%CI)** |
| --- | --- | --- | --- | --- |
| Male | 8 | Thinness | 58 | 124.8 (124, 127) |
| Male | 8 | Normal BMI | 1017 | 128 (127.5, 128.5) |
| Male | 8 | Overweight | 812 | 130 (130, 131) |
| Male | 8 | Obesity | 819 | 131.8 (131.2, 132) |
| Male | 9 | Thinness | 69 | 131 (129.2, 132.5) |
| Male | 9 | Normal BMI | 1228 | 133 (132.4, 133) |
| Male | 9 | Overweight | 991 | 135 (135, 135.5) |
| Male | 9 | Obesity | 951 | 137 (136.6, 137.5) |
| Male | 10 | Thinness | 90 | 134.8 (133.3, 136) |
| Male | 10 | Normal BMI | 1587 | 138 (137.5, 138) |
| Male | 10 | Overweight | 1248 | 140.5 (140, 141) |
| Male | 10 | Obesity | 1217 | 143 (142.7, 143.3) |
| Male | 11 | Thinness | 132 | 140.5 (139.7, 142) |
| Male | 11 | Normal BMI | 2434 | 143.7 (143.3, 144) |
| Male | 11 | Overweight | 1954 | 146 (146, 146.5) |
| Male | 11 | Obesity | 1890 | 148.8 (148.2, 149) |
| Male | 12 | Thinness | 89 | 146 (144.5, 147) |
| Male | 12 | Normal BMI | 1552 | 150.2 (150, 151) |
| Male | 12 | Overweight | 1331 | 153.5 (153, 154) |
| Male | 12 | Obesity | 1261 | 156 (155.5, 156.3) |
| Male | 13 | Thinness | 61 | 155 (152.2, 159) |
| Male | 13 | Normal BMI | 1186 | 159 (158.5, 160) |
| Male | 13 | Overweight | 1026 | 162 (161.2, 162) |
| Male | 13 | Obesity | 978 | 163 (162.8, 164) |
| Male | 14 | Thinness | 35 | 164 (159, 167.5) |
| Male | 14 | Normal BMI | 875 | 165.5 (165, 166) |
| Male | 14 | Overweight | 747 | 167 (166, 167) |
| Male | 14 | Obesity | 697 | 168.5 (167.7, 169) |
| Male | 15 | Thinness | 13 | 170.1 (164.4, 175) |
| Male | 15 | Normal BMI | 365 | 169.3 (169, 170) |
| Male | 15 | Overweight | 325 | 169.8 (169, 170) |
| Male | 15 | Obesity | 299 | 170.5 (170, 171) |
| Male | 16 | Thinness | 13 | 173 (167.4, 181) |
| Male | 16 | Normal BMI | 291 | 171.7 (171, 172.5) |
| Male | 16 | Overweight | 260 | 171 (170, 172) |
| Male | 16 | Obesity | 232 | 172.5 (171.5, 173.4) |
| Male | 17 | Thinness | 4 | 176.5 (-Inf, Inf) |
| Male | 17 | Normal BMI | 82 | 173 (171, 175) |
| Male | 17 | Overweight | 65 | 173.5 (172, 175) |
| Male | 17 | Obesity | 56 | 174.2 (173, 176.5) |
| Male | 18 | Normal BMI | 19 | 176.5 (171.5, 180.5) |
| Male | 18 | Overweight | 16 | 171 (168.5, 174) |
| Male | 18 | Obesity | 13 | 174 (170.5, 179) |
| Female | 7 | Thinness | 133 | 119 (117.6, 120.5) |
| Female | 7 | Normal BMI | 2144 | 121 (121, 121) |
| Female | 7 | Overweight | 851 | 123 (123, 124) |
| Female | 7 | Obesity | 342 | 124 (123.2, 125) |
| Female | 8 | Thinness | 160 | 123.4 (122, 125) |
| Female | 8 | Normal BMI | 2736 | 126.5 (126, 127) |
| Female | 8 | Overweight | 1097 | 129.5 (129, 130) |
| Female | 8 | Obesity | 427 | 130.4 (130, 131) |
| Female | 9 | Thinness | 243 | 130 (129.4, 131.7) |
| Female | 9 | Normal BMI | 4036 | 133 (132.5, 133) |
| Female | 9 | Overweight | 1604 | 135.5 (135, 136) |
| Female | 9 | Obesity | 630 | 137 (136.2, 137.3) |
| Female | 10 | Thinness | 166 | 136 (134.5, 137.8) |
| Female | 10 | Normal BMI | 2601 | 140 (139.5, 140) |
| Female | 10 | Overweight | 1055 | 143 (142.5, 143.3) |
| Female | 10 | Obesity | 418 | 144 (143.3, 145) |
| Female | 11 | Thinness | 100 | 144.1 (142, 146) |
| Female | 11 | Normal BMI | 1839 | 147 (146.7, 147.5) |
| Female | 11 | Overweight | 779 | 149.3 (149, 150) |
| Female | 11 | Obesity | 306 | 150.7 (150, 151.7) |
| Female | 12 | Thinness | 81 | 150.5 (149, 152.5) |
| Female | 12 | Normal BMI | 1381 | 152.3 (152, 153) |
| Female | 12 | Overweight | 574 | 154 (153.5, 154.5) |
| Female | 12 | Obesity | 230 | 155 (154, 156) |
| Female | 13 | Thinness | 31 | 155 (150.6, 158.5) |
| Female | 13 | Normal BMI | 631 | 155.5 (155, 156) |
| Female | 13 | Overweight | 300 | 157 (156.2, 158) |
| Female | 13 | Obesity | 110 | 157.7 (156, 159) |
| Female | 14 | Thinness | 30 | 158 (153.5, 159.5) |
| Female | 14 | Normal BMI | 554 | 157.5 (156.8, 158) |
| Female | 14 | Overweight | 256 | 159 (158, 160) |
| Female | 14 | Obesity | 90 | 159 (158, 160) |
| Female | 15 | Thinness | 6 | 158 (148.5, 168.5) |
| Female | 15 | Normal BMI | 108 | 159.2 (158, 161.5) |
| Female | 15 | Overweight | 50 | 159.5 (157, 161) |
| Female | 15 | Obesity | 26 | 160.2 (156, 162) |
| Female | 16 | Thinness | 6 | 162.2 (155, 167.5) |
| Female | 16 | Normal BMI | 52 | 160 (158, 163) |
| Female | 16 | Overweight | 25 | 160 (155, 162) |
| Female | 16 | Obesity | 6 | 158.2 (154, 171) |

95%CI: 95% confidence interval.
